# COMPARATIVE PERFORMANCE OF TWO AUTOMATED MACHINE LEARNING PLATFORMS FOR COVID-19 DETECTION BY MALDI-TOF-MS

**DOI:** 10.1101/2022.02.02.22270298

**Authors:** Hooman H. Rashidi, John Pepper, Taylor Howard, Karina Klein, Larissa May, Samer Albahra, Brett Phinney, Michelle R. Salemi, Nam K. Tran

**Author notes:** Correspondence [Note: Both authors below are corresponding authors] Nam K. Tran, PhD, HCLD (ABB), FAACC, Professor, Dept. of Pathology and Laboratory Medicine, University of California Davis, 4400 V St., Sacramento, CA 95817, Hooman H. Rashidi MD, MS, FCAP, FASCP, Professor, Dept. of Pathology and Laboratory Medicine, University of California Davis, 4400 V St., Sacramento, CA 95817.

## Abstract

The 2019 novel coronavirus infectious disease (COVID-19) pandemic has resulted in an unsustainable need for diagnostic tests. Currently, molecular tests are the accepted standard for the detection of SARS-CoV-2. Mass spectrometry (MS) enhanced by machine learning (ML) has recently been postulated to serve as a rapid, high-throughput, and low-cost alternative to molecular methods. Automated ML is a novel approach that could move mass spectrometry techniques beyond the confines of traditional laboratory settings. However, it remains unknown how different automated ML platforms perform for COVID-19 MS analysis. To this end, the goal of our study is to compare algorithms produced by two commercial automated ML platforms (Platforms A and B). Our study consisted of MS data derived from 361 subjects with molecular confirmation of COVID-19 status including SARS-CoV-2 variants. The top optimized ML model with respect to positive percent agreement (PPA) within Platforms A and B exhibited an accuracy of 94.9%, PPA of 100%, negative percent agreement (NPA) of 93%, and an accuracy of 91.8%, PPA of 100%, and NPA of 89%, respectively. These results illustrate the MS method’s robustness against SARS-CoV-2 variants and highlight similarities and differences in automated ML platforms in producing optimal predictive algorithms for a given dataset.

## INTRODUCTION

The 2019 novel coronavirus infectious disease (COVID-19) pandemic caused by severe acute respiratory syndrome (SARS) coronavirus (CoV) – 2 has severely disrupted infectious disease testing worldwide.^1^ With the increasing spread of highly infectious SARS-CoV-2 variants (*e*.*g*., Delta, Lambda, Omicron) and challenges in achieving herd immunity in both developed and undeveloped countries, the demand for COVID-19 testing remains exceptionally high for containment as well as to re-open schools and businesses.^2^ Matrix assisted laser desorption ionization (MALDI) – time-of-flight (TOF) – mass spectrometry (MS) serves as a unique alternative COVID-19 screening method that overcomes trade-offs found with commercial molecular and antigen tests (**Figure 1**).^3-5^ MALDI-TOF-MS detects a spectrum of ionizable host and perhaps viral proteins from nasal swab samples. The simple pre-analytic requirements of MALDI-TOF-MS enable it to be rapid, cost-effective, and not reliant on supply chains impacted by the pandemic.

**Fig 1.**
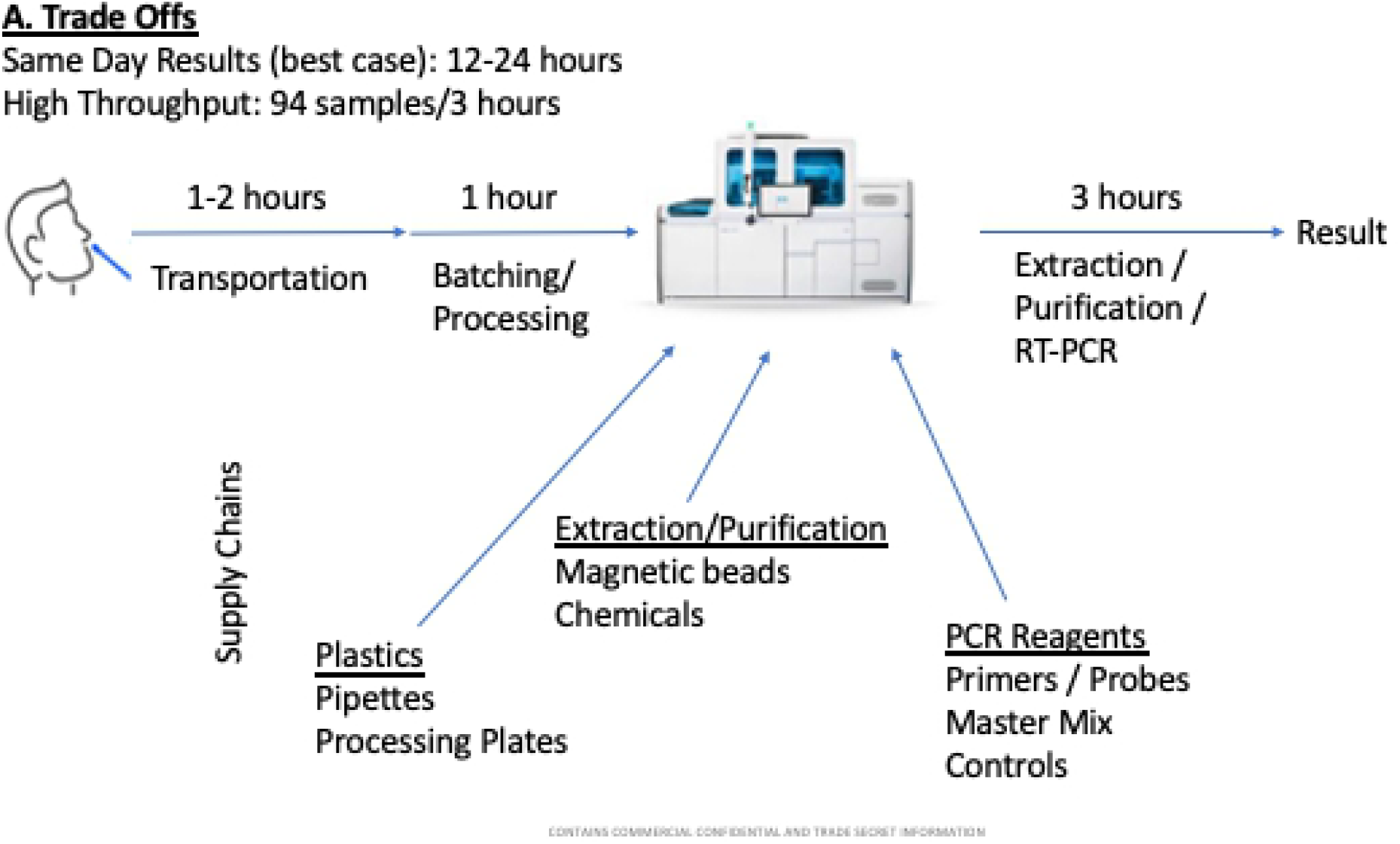
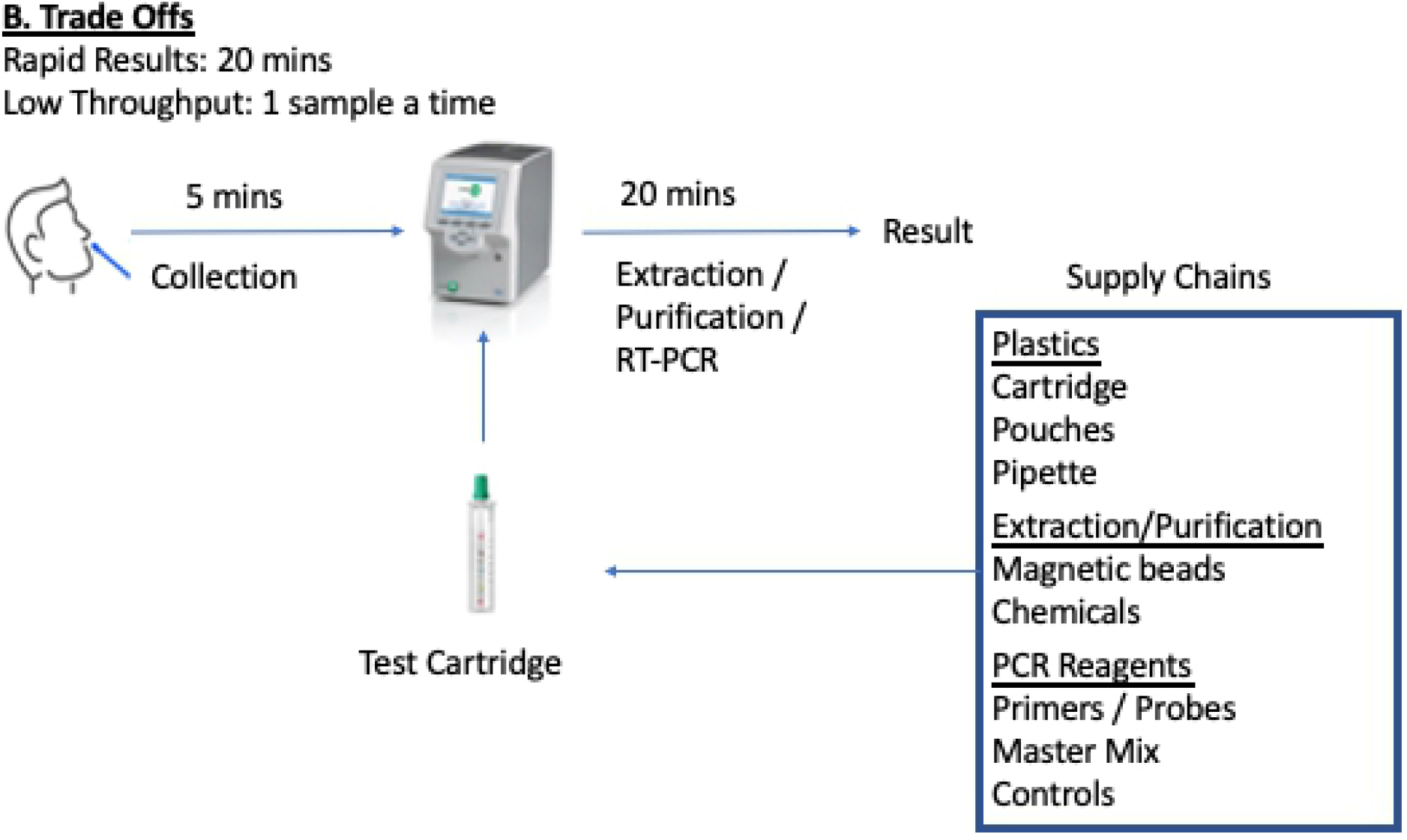
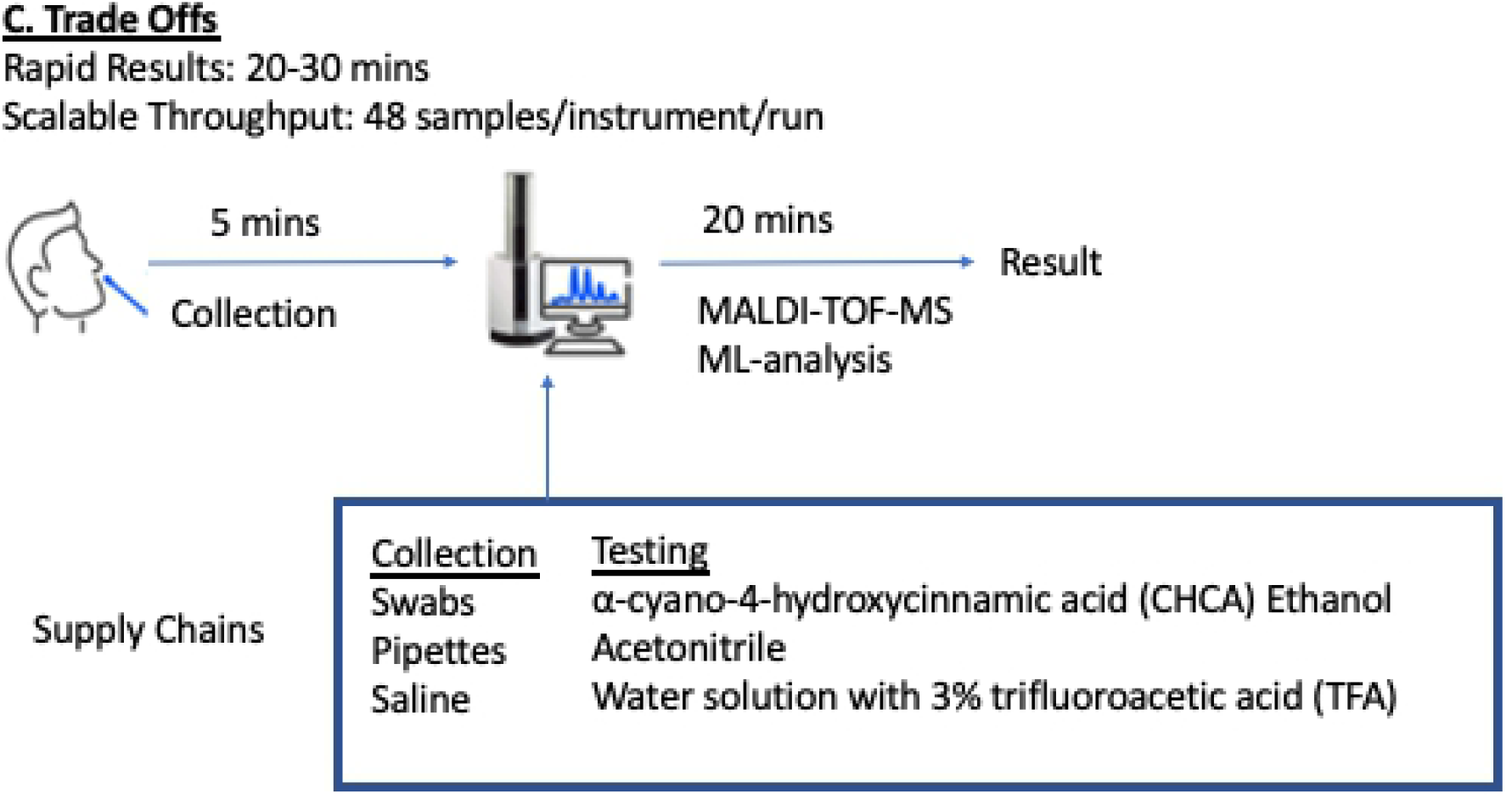
Comparison of Molecular, Antigen, and ML-Enhanced MALDI-TOF-MS Workflows. The figure illustrates the tradeoffs of (A) high throughput molecular platforms, (B) point-of-care molecular platforms, and (C) our proposed MALDI-TOF-MS method.

Interestingly, the sensitivity and specificity for MALDI-TOF-MS when detecting COVID-19 in nasal swab samples is impacted by the shear complexity of the specimen matrix.^3^ Studies suggest that over 1,000 protein groups may be represented in nasopharyngeal (NP) swab specimens when measured by MS techniques.^6^ As such, the differentiation of mass spectra between disease states is not feasible by the human eye – creating a unique opportunity for artificial intelligence (AI) / machine learning (ML) to process and to identify patterns within such complex spectra to enhance their distinguishing performance capabilities.

Traditional ML development tools can be laborious and require significant data science expertise.^7^ Data scientists may also unintentionally prefer one ML technique versus another by nature of their experience, preference, and other confounding factors.^8,9^ Due to these barriers, development of ML models via traditional programming limits the number of models that could be evaluated. The employment of automated ML platforms overcomes many of these challenges and provides means to quickly develop, train, validate, and implement a range of candidate algorithms that span the spectrum of ML techniques.

Recent studies have shown MALDI-TOF-MS enhanced by ML performs with adequate sensitivity and specificity for detecting COVID-19 positive cases.^3,4^ The study by Tran *et al*. highlights the use of ML enhanced MALDI-TOF-MS detection of COVID-19.^3^ Uniquely, this study was the first to use automated ML (AutoML) to rapidly produce algorithms to facilitate MALDI-TOF-MS COVID-19 detection in both symptomatic and asymptomatic patients. However, like with any AI approach, best practices dictate thorough validation across a diverse dataset, and ideally, comparative studies against other ML platforms.^10^ To this end, the goal of this study is to compare the predictive performance of two known AutoML platforms when analyzing MALDI-TOF-MS for COVID-19 positive versus negative individuals.

## MATERIALS AND METHODS

We conducted a retrospective study analyzing data from a COVID-19 clinical trial comparing a MALDI-TOF-MS based approach against reverse transcription (RT) – polymerase chain reaction (PCR) (**Figure 2**). The MALDI-TOF-MS dataset served as the basis for comparing the accuracy along with the positive percent agreement (PPA) and negative percent agreement (NPA) within the two automated ML platforms for detecting COVID-19. Positive percent agreement and NPA terminology is used instead of clinical sensitivity and specificity in these studies to align with United States Food and Drug Administration (FDA) guidelines when evaluating tests that lack a “gold standard” or when an imperfect comparative method exists, such as in the case of COVID-19 detection.^11^

**Fig 2.**
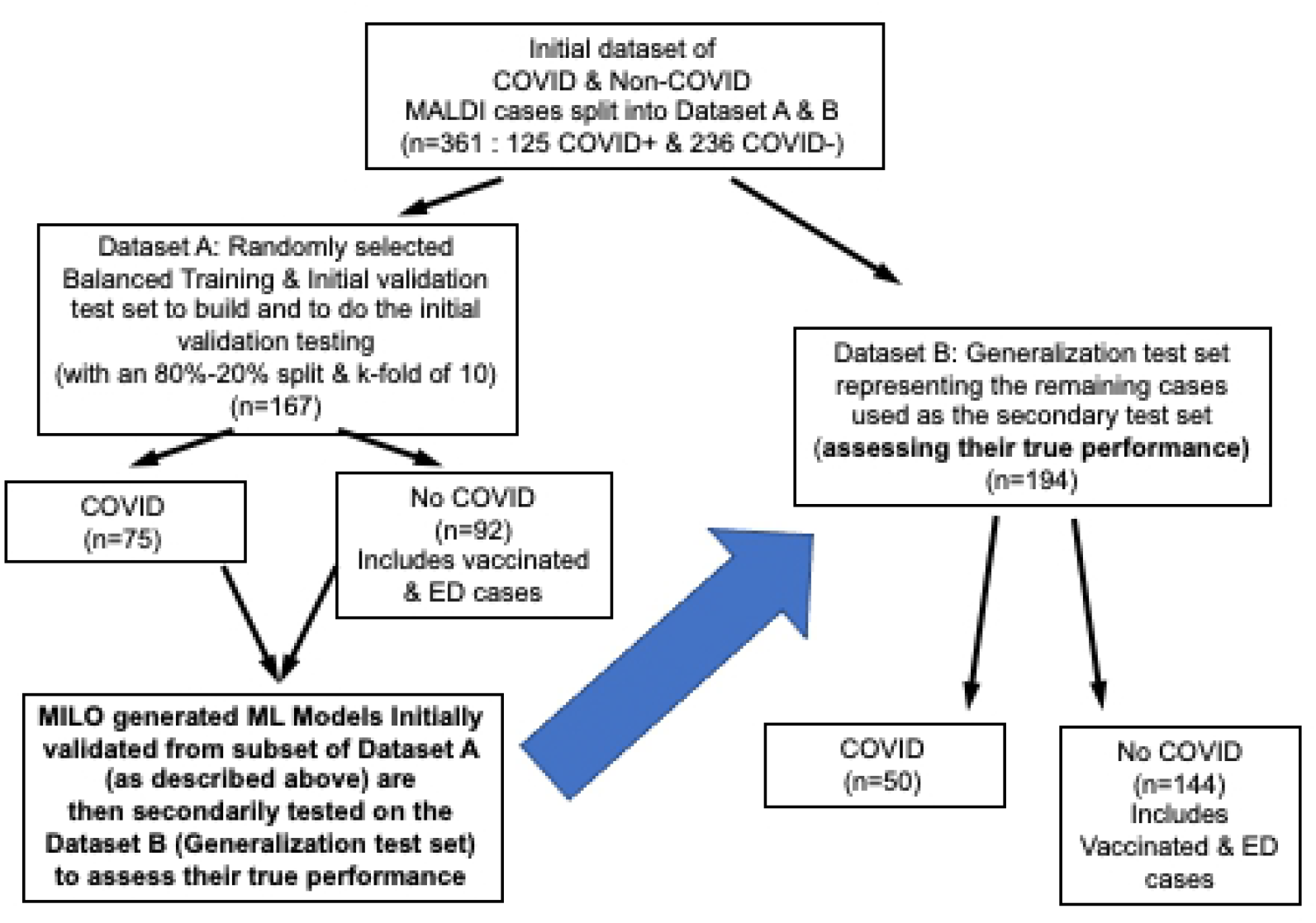
Study Datasets. A total of 361 asymptomatic and symptomatic patients were enrolled. These data were divided into Datasets A and B, with Dataset A serving as the training/ initial validation dataset. Optimized models produced from Dataset A were then secondarily tested with Dataset B for generalization to assess their true performance. Notably, Dataset A and B both contained random samples of the various negative subgroups (negative vaccinated individuals, ED patients, etc.).

### Study Population / Samples

This study was approved by the local Institutional Review Board with informed consent obtained for acquiring the anterior nares nasal swab samples. The study dataset consisted of MALDI-TOF-MS spectra from 361 subjects from December 2020 to August 2021. Among the 361 subjects, 199 were from the first phase of our study (Dataset A) which was conducted prior to emergency use authorization (EUA) of the first COVID-19 vaccines.^3^ An additional 162 patients (Dataset B) were added in the second phase to study additional symptomatic populations meeting COVID-19 testing criteria (*i*.*e*., patients who presented with or without symptoms at the time of collection) as well as asymptomatic volunteers obtained from workplace screening and the emergency medicine population. COVID-19 vaccine status was also included for this new asymptomatic population. Anterior nares nasal swab specimens were collected from each patient and preserved in commercial saline-based viral transport media (Thermo Fisher, Waltham, MA). All samples were tested by MALDI-TOF-MS and RT-PCR.

### COVID-19 Sample Testing

Mass spectrometry testing was performed on a Shimadzu 8020 (Shimadzu Scientific Instruments, Columbia, MD) MALDI-TOF-MS analyzer as described previously.^3^ Briefly, our study tested nasal swabs directly plated onto the MALDI-TOF-MS target plate. MALDI-TOF-MS mass range was set to 2,000 – 20,000 Daltons. Residual saline transport media was then tested by RT-PCR using FDA EUA assays. For Dataset B, all samples were tested by an EUA SARS-CoV-2 digital droplet RT-PCR (ddPCR) test (Bio-Rad, Hercules, CA). The use of ddPCR was to leverage the absolute quantitation capability of the platform and determine SARS-CoV-2 viral RNA load. All ddPCR SARS-CoV-2 RNA positive, or discordant (*e*.*g*., MALDI-TOF-MS negative/ddPCR positive) results were also sequenced. Sequencing was performed using a Respiratory Pathogen Identification Panel (RPIP) (Explify, IDbyDNA, Salt Lake City, Utah) via a MiSeq analyzer (Illumina, San Diego, CA). The RPIP can simultaneously detect the RNA and DNA of over 180 bacteria, 50 fungi, and viruses including SARS-CoV-2 variant identification.

### Automated Machine Learning Approaches

Two AutoML platforms analyzed MALDI-TOF-MS spectra: Platform A (Machine Intelligence Learning Optimizer [MILO], MILO ML, LLC, Sacramento, CA), and Platform B (Microsoft ML.NET, Microsoft, Redmond, WA) (**Table 1**).

**Table 1.** Comparison of AutoML Specifications.

|  | **<u>Platform A</u>** | **<u>Platform B</u>** |
| --- | --- | --- |
| Manufacturer (Location) | MILO-ML, LLC (Sacramento, CA) | Microsoft Corporation (Redmond, WA) |
| Interface | GUI | ML.NET |
| Machine Learning Methods | k-NN, GBM, LR, MLP-NN, NB, RF, SVM | AP-NN, BDT, LBFGS, LGBM, linear SVM, LR,<br>SDCA, SGD |
| Automated Data Feature Selector | ANOVA F select percentile feature selector and RF<br>Feature Importances Selector | None |

**Abbreviations:** ANOVA, analysis of variance; AP, averaged perceptron; BDT, boosted decision tree; GBM, gradient boosting machine; GUI, graphical user interface; k-NN, k-nearest neighbors; LBFGS, limited memory Broyden-Fletcher-Goldfarb-Sanno; LGBM, lightGBM; LR, logistic regression; MLP, multilayer-perceptron; NB, naïve Bayes; NN, neural network; RF, random forest; SDCA, stochastic dual coordinate ascent; SGD, stochastic gradient descent; SVM, support vector machine.

#### Platform A (Machine Intelligence Learning Optimizer)

Briefly, Platform A was also used in the original study, as the AutoML software combines various unsupervised and supervised methods to ultimately acquire the optimized ML model. These include a data processor, data feature selector (ANOVA F select percentile feature selector and RF Feature Importances Selector) and feature set transformer (*e*.*g*., principal component analysis) along with various custom hyperparameter search tools (*i*.*e*., its custom grid search along with its random search tools) within multiple supervised embedded algorithms (*i*.*e*., multi-layer perceptron neural network [NN], logistic regression [LR], naïve Bayes [NB], k-nearest neighbor [*k-*NN], support vector machine [SVM], random forest [RF], and XGBoost gradient boosting machine GBM]). This platform ultimately identifies the optimal hyperparameter combinations within these embedded supervised algorithms/methods to build and analyze thousands of unique optimized models that are then statistically assessed to identify the best performing model for one’s given task.

Within this study, the trial data was imported into Platform A with the outcome goal of acquiring and evaluating the MS peak patterns observed that can then assess COVID-19 status as its final outcome measure. Once the datasets are uploaded into the MILO platform and the target of interest is selected (COVID-19 status in this case), MILO then automatically performs the unsupervised and supervised methods mentioned above. It starts with using a relatively balanced dataset (Dataset A) to train and initially validate the models on an 80%-20% split (80% used to train and 20% used to perform the initial validation of the trained model) with an embedded 10 *k*-fold cross validation. However, the final performance measure of each of the previously described models is not based on the 20% data subsets in Dataset A but rather on a separate secondary/generalization dataset (Dataset B) that has not been part of any of the training or initial validation measures mentioned. This study design approach will minimize the possibility of overfitted ML models and increase the likelihood of acquiring more generalizable ML models, especially in studies that are based on smaller datasets. In addition, the platform also makes no assumptions about any of the unsupervised and more importantly the supervised ML methods employed. Its ultimate function is to create a large set of combinations from these essential ML elements. Hence, ultimately creating a large number of ML pipelines (*i*.*e*., combination of various scalers, scorers, various unsupervised feature selectors/transformers, hyperparameter searcher tools, and supervised algorithms) to generate thousands of optimized ML models each with potentially different unique characteristics and performance capabilities (*e*.*g*., some optimizing sensitivity while others optimizing specificity or other performance measures such as but not limited to PPV, NPV, F1 score, and Brier score) which ultimately translates to broader ML model selection options for the investigator in their given task.

#### Platform B (Microsoft ML.NET)

Platform B is an open source, cross platform, ML framework with an automated hyperparameter search functionality in the form of AutoML.^12^ The platform is designed to facilitate the embedding of machine learning in .NET applications. Platform B utilizes a similar set of available supervised algorithms and functionality but without any unsupervised embedded steps. Platform B functionality is available through the ML.NET package with a .NET language, through Python with NimbusML, with a graphical user interface as part of Visual Studio 2019 using Model Builder, or through a command line interface. In order to duplicate the precise generalization dataset strategy employed by Platform A, the authors used the .NET package and a customization of the binary classification AutoML example available on the Platform B samples via GitHub (San Francisco, CA). As noted, unlike Platform A, Platform B does not perform automated feature selection or transformation but only hyperparameter search using SMAC. Platform B begins by running all available trainers in with defaults and selecting the top three. Then each of the three is run for 20 iterations followed by Bayesian optimization. The algorithms supported by Platform B include averaged perceptron NN, LR optimized with stochastic dual coordinate ascent (SDCA), stochastic gradient descent (SGD), symbolic SGD, or limited memory Broyden-Fletcher-Goldfarb-Sanno (LBFGS), linear SVM, FastTree Multiple Additive Regression Trees (MART) GBM, FastForest RF implementation, and light gradient boosting machines (LGBM). Platform B also includes other models including a non-linear SVM and a generalized additive model, but these were currently unavailable for hyperparameter optimization as the AutoML component is focused on an efficient search space. However, Platform B does allow the user to limit search to only a specific subset of the algorithms which was used in this project to determine the best hyperparameters for those models outside the top three.

### Statistical Analysis

JMP software (SAS, Cary, NC) was used for traditional statistical analyses. Descriptive statistics were used to compare viral RNA load between study populations. Continuous parametric independent data was compared by the 2-sample *t-*test.

## RESULTS

### Study Population

A total of 361 subjects were enrolled and tested by both MALDI-TOF-MS and RT-PCR (**Figure 2**). From the 361 samples tested, 125 samples were COVID-19 positive with 236 determined to be negative (**Table 2**). For positive individuals, SARS-CoV-2 variants included 49.6% Alpha (B.1.1.7), 0.8% Gamma (P.1), 16.8% Delta (B.1.617), 8% Iota (B.1.526), 16.8% B.1.1, and 8% B.1.168. Within the 236 negative cases mentioned, 45 were symptomatic including 3 patients who were positive by sequencing (1 patient with human coronavirus NL63, and 2 patients with *Staphylococcus aureus* respectively). One-hundred ninety-one COVID-19 negative patients were asymptomatic with 74 being recently vaccinated with EUA mRNA-based vaccines.

**Table 2.** Study Population.

|  | **<u>COVID-19 Positive (n = 125)</u>** | **<u>COVID-19 Negative (n = 236)</u>** | **<u>P-Value</u>** |
| --- | --- | --- | --- |
| Mean (SD) Age (Years) | 42.7 (15.3) | 40.9 (16.1) | NS |
| Percent Symptomatic | 88.9% (111/125) | N/A | <0.001 |
| Percent Vaccinated | 0% (0/125) | 31.4% (74/236) | <0.001 |
| Mean (SD) Viral RNA Load (copies/mL) |  |  |  |
| *N1 Target:* | 37,549.7 (19,236.2) | N/A | N/A |
| *N2 Target:* | 779.0 (237.3) | N/A | N/A |

**Abbreviations:** COVID-19, novel coronavirus infectious disease 2019; N/A, not applicable; NS, not significant; RNA, ribonucleic acid; SD, standard deviation

As noted in **Figure 1**, two randomly generated datasets were acquired for the ML studies from the total 361 cases, designated as the training/initial validation dataset (Dataset A) and the secondary/generalization dataset (Dataset B). Dataset A (used for the ML training and its initial validation) is a relatively balanced dataset of 167 cases containing a randomly selected 75 COVID positive cases and 92 negative cases. The 92 negative cases within dataset A contained 30 of the vaccinated samples, 20 of the ED cases, the 1 non-COVID-19 viral case, 1 of the 2 bacterial cases and 40 of other asymptomatic negative samples. The remaining cases comprised Dataset B (secondary / generalization dataset) which contained 50 COVID positive cases and 144 negative cases. The 144 negative cases within dataset B contained 44 of the vaccinated samples, 25 of the ED cases, the 1 of the 2 bacterial cases and 74 of the other asymptomatic negative samples.

### Data Analysis & Machine Learning

Using the two AutoML platforms, ML analysis was conducted to build and determine the best performing model for the given task of distinguishing the COVID positive cases from the negative cases within the dataset mentioned above with the goal of selecting and optimizing the model’s PPA (*i*.*e*., sensitivity). Each AutoML platform was fed the same two datasets (Dataset A and Dataset B) as detailed above and followed the same study approach. As mentioned, Dataset A is used for the model training / initial validation steps while Dataset B is used solely as the hold-out dataset for secondary/generalization testing of the models. Dataset A is a relatively balanced dataset with 167 total cases (75 COVID positive cases and 92 negative cases). The 92 negative cases within dataset A contain 30 of the vaccinated samples, 20 of the ED cases, the 1 non-COVID-19 viral case, 1 of the 2 bacterial cases and 40 of other asymptomatic negative samples. Dataset B (secondary/generalization dataset) contained 50 COVID-19 positive cases and 144 negative cases. The 144 negative cases within dataset B contained 44 of the vaccinated samples, 25 of the ED cases, the 1 of the 2 bacterial cases and 74 of the other asymptomatic negative samples.

Platform A’s AutoML software was trained and initially validated on Dataset A with an 80-20 split and 10 k-fold cross validation which generated 400,869 models which were then secondarily tested on Dataset B to assess the model’s true performance measures (**Table 3**). The process was completed in approximately 5.5 hours using a high-performance platform with a multi-core personal computer. With the goal of maximizing the PPA, the following 2 models were identified: the first is a NN model using all features that exhibited an accuracy of 94.9% (95% CI, 90.7-97.5), PPA of 100% (95% CI, 92.9-100), NPA of 93% (95% CI, 87.6-96.6), followed by a support vector machine model using 75% of the features showing an accuracy of 93.3% (95% CI, 88.8-96.4), PPA of 100% (95% CI, 92.9-100), and NPA of 91% (95% CI, 85.1-95.1).The best two models from Platform B were found to be an SDCA-LR model with an accuracy of 91.8% (95% CI, 86.9-95.2), PPA of 100% (95% CI, 92.9-100), and NPA of 89% (95% CI, 82.6-93.5) and a SVM model with an accuracy of 95.4% (95% CI, 91.4-97.9), PPA of 98% (95% CI, 89.4-99.9), and NPA of 94% (95% CI, 89.4-97.6), both using all the features (i.e. all MALDI-TOF-MS peaks; **Table 3**).

**Table 3.** Machine Learning Algorithm Generalization Performance for Top Models Produced by Platforms A and B.

| **A. MILO AutoML generated Models** |  |  |  |  |  |  |
| --- | --- | --- | --- | --- | --- | --- |
| **Method** | **Accuracy %**<br>**(95% CI)** | **AUROC**<br>**(95% CI)** | **Positive Percent**<br>**Agreement (PPA)**<br>**% (95% CI)** | **Negative Percent**<br>**Agreement (NPA)**<br>**% (95% CI)** | **F1 Score** | **% features**<br>**Selected** |
| LBFGS-Logistic Regression | 92.8 (88.2-96.0) | 98.9 (81.9-100) | 100 (92.9-100) | 90.3 (84.2-94.6) | 91.3 | All* |
| k-Nearest Neighbor | 92.3 (87.6-95.6) | 96.9 (60.1-100) | 100 (92.9-100) | 89.6 (83.4-94.1) | 90.7 | 25%<sup>#</sup> |
| Naïve Bayes | 91.7 (86.9-95.2) | 99.2 (84.8-100) | 100 (92.9-100) | 88.9 (82.6-93.5) | 90.2 | All* |
| Random Forest | 95.4 (91.4-97.9) | 98.1 (83.3-100) | 92.0 (80.8-97.7) | 96.5 (92.1-98.9) | 93.9 | All* |
| Support Vector Machine | 93.3 (88.8-96.4) | 98.6 (86.8-100) | 100 (92.9-100) | 91.0 (85.1-95.1) | 91.9 | 75%<sup>##</sup> |
| Neural Network-Multi Layer Perceptron | 94.9 (90.7-97.5) | 99.6 (84.9-100) | 100 (92.9-100) | 93.1 (87.6-96.6) | 92.5 | All* |
| Gradient Boosting Machine (XGBoost) | 93.8 (89.4-96.8) | 98.3 (82.0-100) | 94.0 (83.5-98.7) | 93.8 (88.5-97.1) | 92.2 | All* |

| **B. Microsoft AutoML generated Models** |  |  |  |  |  |  |
| --- | --- | --- | --- | --- | --- | --- |
| **Method** | **Accuracy %**<br>**(95% CI)** | **AUROC \*\*** | **Positive Percent**<br>**Agreement (PPA)**<br>**% (95% CI)** | **Negative Percent**<br>**Agreement (NPA)**<br>**% (95% CI)** | **F1 Score** | **% features**<br>**Selected** |
| Fast Tree | 87.1 (81.6-91.5) | 98.0 | 98.0 (89.4-99.9) | 83.3 (76.2-89.0) | 79.7 | All |
| Fast Forest | 86.6 (80.9-91.1) | 96.9 | 92.0 (80.8-97.8) | 84.7 (77.8-90.2) | 78.0 | All |
| Gradient Boosting Machine (light) | 86.1 (80.4-90.6) | 98.3 | 98.0 (89.4-99.9) | 81.9 (74.7-87.9) | 78.4 | All |
| Support Vector Machine | 95.4 (91.4-97.9) | 99.5 | 98.0 (89.4-99.9) | 94.4 (89.4-97.6) | 91.6 | All |
| SDCA-Logistic Regression | 91.8 (86.9-95.2) | 99.4 | 100 (92.9-100) | 88.9 (82.6-93.5) | 86.2 | All |
| LBFGS-Logistic Regression | 90.7 (85.7-94.4) | 99.3 | 100 (92.9-100) | 87.5 (80.9-92.4) | 84.8 | All |
| SGD-Calibrated | 91.2 (86.3-94.8) | 99.1 | 98.0 (89.4-99.9) | 88.9 (82.6-93.5) | 85.2 | All |
| Symbolic SGD-Logistic Regression | 85.6 (79.8-90.2) | 97.1 | 92.0 (80.8-97.8) | 83.3 (76.2-89.0) | 76.7 | All |
| Averaged Perceptron | 89.2 (83.9-93.2) | 98.7 | 98.0 (89.4-99.9) | 86.1 (79.4-91.3) | 82.4 | All |

\*\*95% CI is not reported by the Microsoft AutoML platform for the calculated AUROC
\*All features were used on a PCA transformed data

## DISCUSSION

As the COVID-19 pandemic continues across the globe with highly populous but less equipped (*e*.*g*., healthcare access, vaccine availability, testing capacity) countries (*e*.*g*., India and Latin America)^13,14^ being devastated by the new surges, and simultaneously, as COVID-19 reservoirs continue to persist and new variants arise due to vaccine hesitancy observed in more well-resourced nations^15-17^, alternatives to costly and resource constrained molecular tests remain critically needed. To this end, due to the continued threat of COVID-19, the global community requires a rapid, cost-effective, and high throughput testing approach to limit transmission of COVID-19.^18^ Molecular methods such as RT-PCR are known to be very sensitive and specific, however, high throughput platforms perform testing in batches—requiring hours or days to produce results.^3,19^ Ultra-fast RT-PCR methods are now available including at the point of care; however, these platforms are limited to testing one sample at a time.^19^ Antigen approaches provide a low-cost alternative to molecular methods; however, sensitivity and specificity suffers, especially for asymptomatic populations.^20^ The use of AutoML with MALDI-TOF-MS for COVID-19 screening is an innovative rapid *and* high-throughput approach at overcoming limitations inherent with molecular- or antigen-based approaches (**Tran 2021**).

However, ML algorithms are only as good as the quality of the data and programmers.^3,8,10,21-23^ Quality of data can be controlled by study design; however, the validity of ML algorithms must be rigorously scrutinized especially for health care applications.^8,9^ Traditional approaches developing ML models relies on expert data scientists to program, and lengthy experimentation cycles to select optimal features for training-testing. Due to the laborious nature of programming, data scientists may select features and limit model development to ML techniques based on their “expertise” and “familiarity” – creating a potential source of bias.

Automated ML platforms such as MILO and Microsoft ML.NET provide means to accelerate development of ML algorithms. These AutoML platforms can go through a much higher number of combination of features across a range of ML techniques in a matter of hours or days. However, different AutoML platforms employ different functions which can influence model development. To this end, the comparison AutoML platforms are necessary to verify performance especially for medical applications.

To our knowledge, this is the first study comparing AutoML platforms performance for COVID-19 screening. The study observed differences in models produced by Platforms A versus B. Although the two Auto-ML platforms showed many similarities in performance measures, the Platform A was able to identify models with greater accuracy, PPA and NPA compared to Platform B. Objectively, Platform A also incorporated unsupervised feature selection methods which were not available on Platform B, which could explain part of the performance enhancements noted on Platform A. Furthermore, this study also reports performance of the ML-enhanced MALDI-TOF-MS method with a larger and more diverse sample size including vaccinated versus unvaccinated, symptomatic versus asymptomatic individuals, and a range of SARS-CoV-2 viral loads as well as COVID-19 variants.

Study limitations include evaluating only two AutoML platforms and a limited sample size. Selection of Platform A was purely by natural extension of our previous study. Comparison in Platform B was based on selecting an AutoML software that was commonly available. We were not able to fully evaluate the performance of this test in non-COVID-19 respiratory infections due to incredibly low prevalence except for one case involving human coronavirus NL63. Likewise, we were not able to evaluate testing performance against the omicron variant due to the enrollment period ending prior to Fall 2021.

## CONCLUSIONS

Validation and reproducibility of the machine learning platforms used to assess and deploy the machine learning component of ML-based testing platforms such as our novel ML-based MALDI-TOF-MS screening tool is an essential step in their overall authenticity confirmation. These comparative studies should be recommended for all testing platforms that employ and incorporate such predictive analytics tools within their testing platform. The concordance of our two AutoML platforms and the robustness of our ML algorithm in the presence of several SARS-CoV-2 variants further supports the future use of such ML-based MALDI-TOF-MS screening tool COVID-19 detection, especially within large gatherings or workplace settings. Further studies looking at non-COVID-19 infectious etiologies are required to further enhance the distinguishing capabilities of our current ML models which would in turn strengthen their true potential and screening capabilities for a variety of infectious agents, including future pandemics.

## Data Availability

Requests should be submitted to UC Davis and SpectraPass investigators. A formal material transfer agreement would need to be set up. Shared data would be limited to de-identified information.

## Acknowledgements

We thank University of California, Davis Health, Ms. Allyson Sage, and Mr. Scott Bainbridge for his support of this study. The project was developed jointly between UC Davis and SpectraPass, LLC.

## Notes

### Competing Interest Statement

N.T. is a consultant for Roche Diagnostics and Roche Molecular Systems. He is also a co-inventor / co-owner of MILO-ML, LLC. H.R. is also a co-inventor / co-owner of MILO-ML, LLC. J.P. is a co-founder and employee of SpectraPass, LLC. L.M. is a consultant for Roche Diagnostics and Roche Molecular Systems. S.A. is a co-inventor / co-owner of MILO-ML, LLC. T.H., K.K., B.P., and M.S. have no competing interests.

### Funding Statement

N.T., H.R., L.M., K.K., T.H., B.P., and M.S. are funded under a sponsored study agreement between UC Davis and SpectraPass, LLC. S.A. received no specific funding for this work. J.P. is an employee of SpectraPass, LLC. SpectraPass, LLC is developing this assay in partnership with UC Davis investigators and were involved with study design, data analysis, decision to publish.

### Author Declarations

The UC Davis Institutional Review Board approved this study.

## REFERENCES

1. World Health Organization, COVID-19 website: https://www.who.int/health-topics/coronavirus#tab=tab_1, Accessed on August 19, 2021.

2. World Health Organization, COVID-19 Variant Tracking website: https://www.who.int/en/activities/tracking-SARS-CoV-2-variants/, Accessed on August 19, 2021.

3. Tran NK, Howard T, Walsh R, et al. Novel Application of Automated Machine Learning With Maldi-Tof-Ms For Rapid High-Throughput Screening of Covid-19. Sci Rep 2021;11:8219.

4. Nachtigall FM, Pereira A, Trofymchuk OS, et al. Detection of SARS-CoV-2 in nasal swabs using MALDI-MS. Nat Biotech 2020;38:1168–1173.

5. Patel R. MALDI-TOF-MS for the diagnosis of infectious diseases. Clin Chem 2015;61:100–111.

6. Gouvia D, Miotello G, Gallais F, et al. Proteotyping SARS-CoV-2 virus from nasopharyngeal swabs: A proof-of-concept focused on a 3 min mass spectrometry window. J Proteom Res 2020;19:4407–4416.

7. Cutillo CM, Sharma KR, Foschini L, et al. Machine intelligence in healthcare— perspectives on trustworthiness, explainability, usability, and transparency. Digital Med 2020;47:1–5.

8. Gianfrancesco MA, Tamang S, Yazdany J, et al. Potential biases in machine learning algorithms using electronic health record data. JAMA Intern Med 2018;178:1544–1547.

9. Wenlong S, Nasraoui O, Shafto P. Evolution and impact of bias in human and machine learning algorithm interaction. PLoS One 2020;15:e0235502.

10. Rashidi HR, Tran NK, Vali Betts E, et al. Artificial Intelligence and Machine Learning in Pathology: The Present Landscape of Supervised Methods. Acad Pathol 2019;6:2374289519873088.

11. Food and Drug Administration Policy for Coronavirus Disease-2019 Tests During the Public Health Emergency (Revised, May 11, 2020): https://www.fda.gov/media/135659/download, Accessed on July 2, 2021.

12. Ahmed Z, Amizadeh S, Bilenko M, et al. Machine learning at Microsoft with ML.NET. In Proceedings of the 25th ACM SIGKDD International Conference on Knowledge Discovery & Data Mining (KDD ‘19). Association for Computing Machinery, New York, NY, USA, 2448–2458. DOI:https://doi.org/10.1145/3292500.3330667.

13. Shivangi, and Meena LS. A comprehensive review of COVID-19 in India: A frequent catch of information. Biotechnol Appl Biochem 2021;12:10.

14. Talha B. COVID-19 in Latin America. Lancet Infect Dis 2020;20:547–548.

15. Wouters OJ, Shadlen KC, Salcher-Konrad M, et al. Challenges in ensuring global access to COVID-19 vaccines: production, affordability, allocation, and development. Lancet 2021;397:1023–1034.

16. Coustasse A, Kimble C, Maxik K. COVID-19 and vaccine hesitancy: A challenge the United States must overcome. J Ambul Care Manage 2021;44:71–75.

17. Sallam M. COVID-19 Vaccine Hesitancy Worldwide: A Concise Systematic Review of Vaccine Acceptance Rates. Vaccines 2021;16:160.

18. Mina M, Larremore DB. COVID-19 test sensitivity – a strategy for containment. N Engl J Med 2020;383:e120.

19. Hanson G, Marino J, Wang ZX, et al. Clinical performance of the point-of-care cobas Liat for detection of SARS-CoV-2 in 20 minutes: A multicenter study. J Clin Microbiol 2020 [in press].

20. Du Z, Pandey A, Bai Y, et al. Comparative cost-effectiveness of SARS-CoV-2 testing strategies in the USA: A modelling study. Lancet 2021;6:E184–E191.

21. Rashidi HH, Makley A, Palmieri TL, et al. Enhancing Military Burn-and Trauma-Related Acute Kidney Injury Prediction Through an Automated Machine Learning Platform and Point-of-Care Testing. Arch Pathol Lab Med. 2021 Mar 1;145(3):320–326.

22. Tran NK, Albahra S, Pham TN, et al. Novel application of an automated-machine learning development tool for predicting burn sepsis: proof of concept. Nature’s Sci Rep. 2020 Jul 23;10(1):12354.

